# Assessment of symptoms, quality of life and remnant gastric activity following gastric bypass using Gastric Alimetry^®^

**DOI:** 10.1101/2023.12.14.23299974

**Authors:** Tim Hsu-Han Wang, Chris Varghese, Stefan Calder, Armen Gharibans, Nicholas Evennett, Grant Beban, Gabriel Schamberg, Greg O’Grady

## Abstract

**Background:** While most gastric bypass patients recover well, some experience long-term complications, including nausea, pain, stricture, and dumping. This study aimed to evaluate symptoms and quality of life (QoL) together with remnant stomach function using the novel Gastric Alimetry® system.

**Method:** Gastric bypass and conversion-to-bypass patients were recruited. The Gastric Alimetry system (Auckland, NZ) was employed, comprising a high-resolution electrode Array, and validated symptom logging App. The protocol comprised 30-minute fasting baseline, a 218kCal meal stimulus, and 4-hours of post-prandial recordings. Symptoms and QoL were evaluated using validated PAGI questionnaires. Remnant gastric electrophysiology evaluation included frequency, BMI-adjusted amplitude, and Gastric Alimetry Rhythm Index (GA-RI; reflecting pacemaker stability), with comparison to matched controls.

**Results:** 38 participants were recruited with mean time from bypass 46.8 ± 28.6 months. One third of patients showed moderate to severe post-prandial symptoms, with patients PAGI-SYM 28 ± 19 vs controls 9 ± 17; PAGI-QoL 37 ± 31 vs 135 ± 22 (p<0.01). Remnant gastric function was markedly degraded shown by undetectable frequencies in 84% (vs 0% in controls), and low GA-RI (0.18 ± 0.08 vs 0.51 ± 0.22 in controls; p<0.0001). Impaired GA-RI and amplitude were correlated with worse PAGI-SYM and PAGI-QOL scores.

**Conclusion:** One third of post-bypass patients suffered significant upper GI symptoms with reduced QoL. The bypassed remnant stomach shows highly deranged electrophysiology, reflecting disuse degeneration. These derangements correlated with QoL, although causality was not addressed.

## Introduction

Gastric bypass is a common procedure that is predominantly performed for obesity, with a long track record of safety and efficacy (1). Sleeve gastrectomy is an alternative procedure, which sometimes may be converted to bypass in the event of intractable complications such as reflux, hiatus hernia and strictures (2).

While varying techniques for bypass exists (e.g., Roux-en-Y, single-anastomosis, mini-loop, omega-loop), all involve the formation of small gastric pouch, connecting the esophagus with the small intestine, and a gastric remnant that is excluded from digestive continuity. The majority of patients do well after these bariatric procedures, however, a proportion experience long-term complication, including nausea, abdominal pain, anastomotic stricture and dumping syndromes (2–4). These can significantly impact quality of life, with evidence suggesting variable return to baseline quality of life dependent on the procedure performed and weight change achieved (5–8).

While there are several studies evaluating symptoms and quality of life after bypass surgery (5–8), meal associations have not been characterized in detail. In addition, the functional physiology of the remnant stomach is largely unknown. It can be expected that a remnant stomach will become dormant and atrophic following the diversion of contents, however, the stomach likely also remains continuously bioelectrically and neurally active, which could be relevant to symptom (9, 10). Gastric dysrhythmias are of interest as they have been implicated in chronic nausea (11–13), as well as post-surgical symptoms after sleeve gastrectomy, esophagectomy, and partial gastrectomy (14–16), albeit exclusively in patients with stomachs still in continuity to date.

Gastric Alimetry® (Alimetry, New Zealand) is a new non-invasive test to evaluate gastric electrophysiology and function at high resolution (HR) (13). The test is extensively validated and has received regulatory approvals for clinical use (9, 15, 17). It also includes a validated evaluation of patient-reported symptom evolution in relation to a meal, using an App, which correlates to quality of life (18). The aim of this study was therefore to assess the symptoms and quality of life (QoL) of patients following gastric bypass and conversion to bypass, as well as remnant gastric function, using the new Gastric Alimetry system.

## Methods

### Patient population

Consecutive patients who underwent either a gastric bypass or a sleeve gastrectomy with subsequent conversion to gastric bypass performed at Auckland City Hospital (Auckland, New Zealand) between 2013-2022 were recruited. Ethics approval was obtained from the Auckland Health Research Ethics Committee (AH1125) and informed consent was obtained from all patients. Patients who had a history of skin allergy, evidence of mechanical gastric or small bowel obstruction as a cause for their symptoms, BMI >40 and those with insulin-dependent diabetes were excluded. Clinical data including operation notes, imaging, endoscopy, and histopathology (if applicable) were evaluated. Patients were also followed up within 1 week for adverse reactions. A healthy control cohort was also recruited, who had a similar caloric content meal, being matched based on patient gender, age and BMI.

### Quality of Life Assessments

At the time of the Gastric Alimetry, validated questionnaires were employed including the PAGI-SYM, PAGI-QOL and EQ-5D-5L which assesses the patients’ symptoms and quality of life over the preceding 2 weeks (19–21). Individual and total scores were obtained and analysed.

### Gastric Alimetry

Gastric Alimetry was performed under a protocol adapted for patients with a reduced gastric volume. This device comprises an HR stretchable electrode array (8×8 electrodes; 20 mm spacing; 196 cm^2^), a wearable Reader, validated iOS app for symptom logging, and a cloud-based reporting platform (**Figure 1A,B**) (18, 22, 23). Baseline recordings were performed in the first 30 minutes, followed by a 218kCal meal, comprising of 100mL of Ensure (93kCal; Abbott Nutrition, IL, USA) and half an oatmeal energy bar (125kcal, 2.5g fat, 22.5g carbohydrate, 5g protein, 3.5g fibre; Clif Bar & Company, CA, USA) to account for the partial loss of stomach reservoir, consumed over 10 minutes and a 4-hr postprandial recording in order to capture a full gastric activity cycle. Patients were seated in a reclined chair and were asked to limit movement, talking, and sleeping, but were able to read, watch media, work on a mobile device, and mobilize for comfort breaks. Symptom development or changes were recorded on the Gastric Alimetry App, calculated into a Total Symptom Burden Score (TSBS) as defined and validated by Sebaratnam *et al* (18).

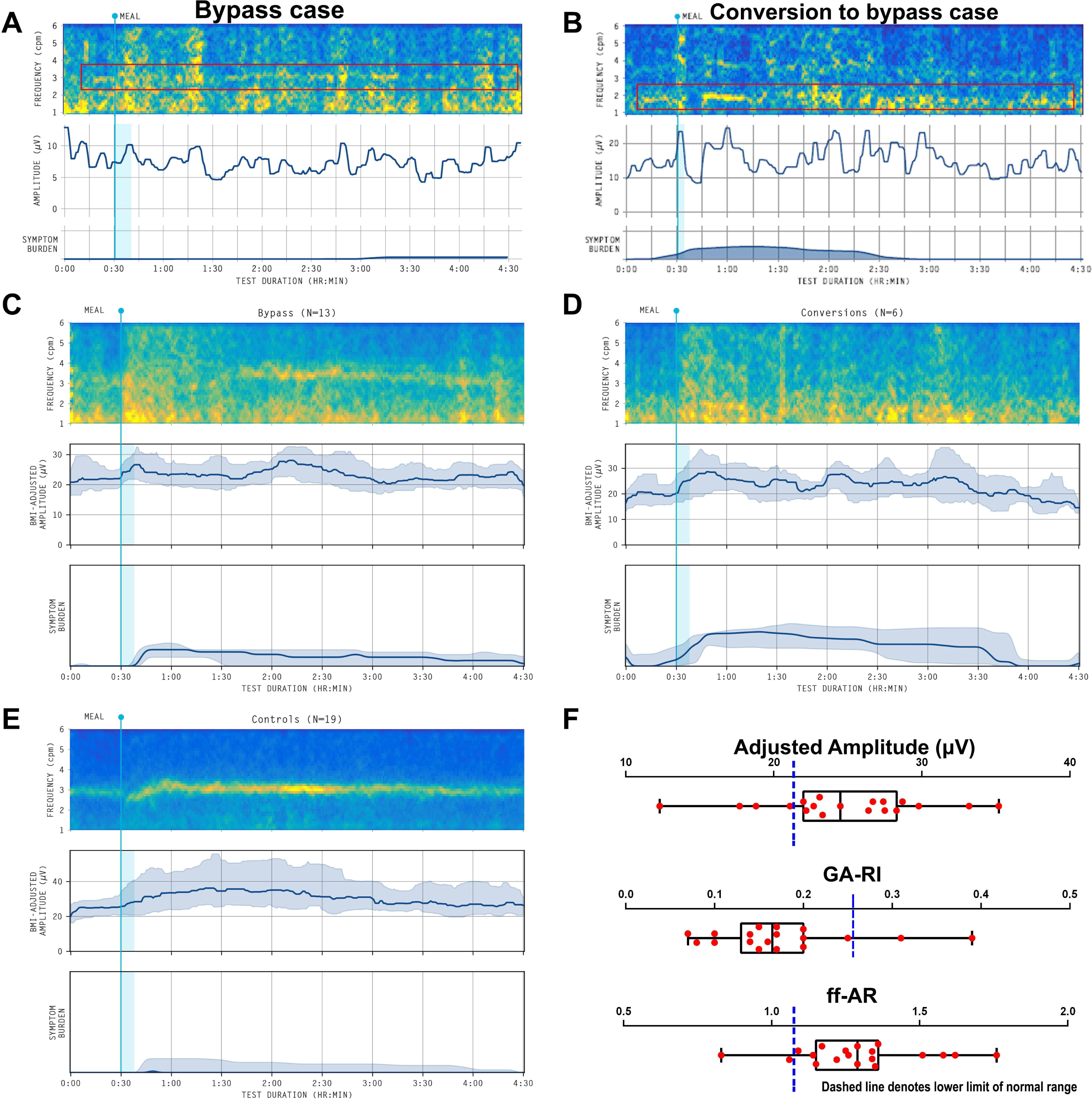

Spectral (frequency-amplitude) analysis was performed, encompassing four established metrics derived from the Gastric Alimetry system (24): Principal Gastric Frequency (PGF), BMI-Adjusted Amplitude, Gastric Alimetry Rhythm Index (GA-RI; reflecting gastric pacemaker activity and stability), fed:fasted Amplitude Ratio (ff-AR; indicating meal response with contractions), with comparison to reference intervals and the matched control cohort separately (25). Frequency was not reported if there was no stable rhythm (as automatically determined in the Gastric Alimetry Report) (23). Adverse events were recorded.

### Data Analyses

Statistical analysis was performed using GraphPad Prism (San Diego, CA, USA) and R v.4.0.1 (R Foundation for Statistical Computing, Vienna, Austria). Analyses were performed on bypass-only cohort, conversion to bypass-only cohort and a combined cohort. Symptom and quality of life comparisons with the matched healthy controls were performed using the unpaired Student’s t-test. Correlation analysis between spectral metrics, TSBS and QoL scores were performed with Spearman’s correlation. All data is presented as mean +/- SD unless otherwise stated, with p<0.05 considered as statistically significant.

## Results

### Patient population

20 patients were recruited in total: 13 bypasses, 6 sleeve converted to bypass, 1 resectional gastric bypass (8 males, 12 females; median age 55.5 years old; age range 33-68 years old; median BMI 30.1; BMI range 21-38.8). Indication for all bypasses were for obesity. Indications for the sleeve converted to bypass included reflux and hiatus hernia (n=5) and narrowing of post-sleeve gastrectomy stomach and twisting (n=1). The resectional gastric bypass was a patient who developed intermittent gastric distention following fundoplication without any evidence of a mechanical cause (gastric pouch but no remnant stomach *in situ*), and was therefore subsequently excluded from the main analysis (data available in **Supplementary** Figure 1). All other reconstructions were performed with a Roux-en-Y reconstruction and no gastric resection. Median time from surgery was 43.5 months (range 4-107 months). Patient and matched control demographics are presented in **Table 1**, showing no differences in BMI, age, or gender between groups.

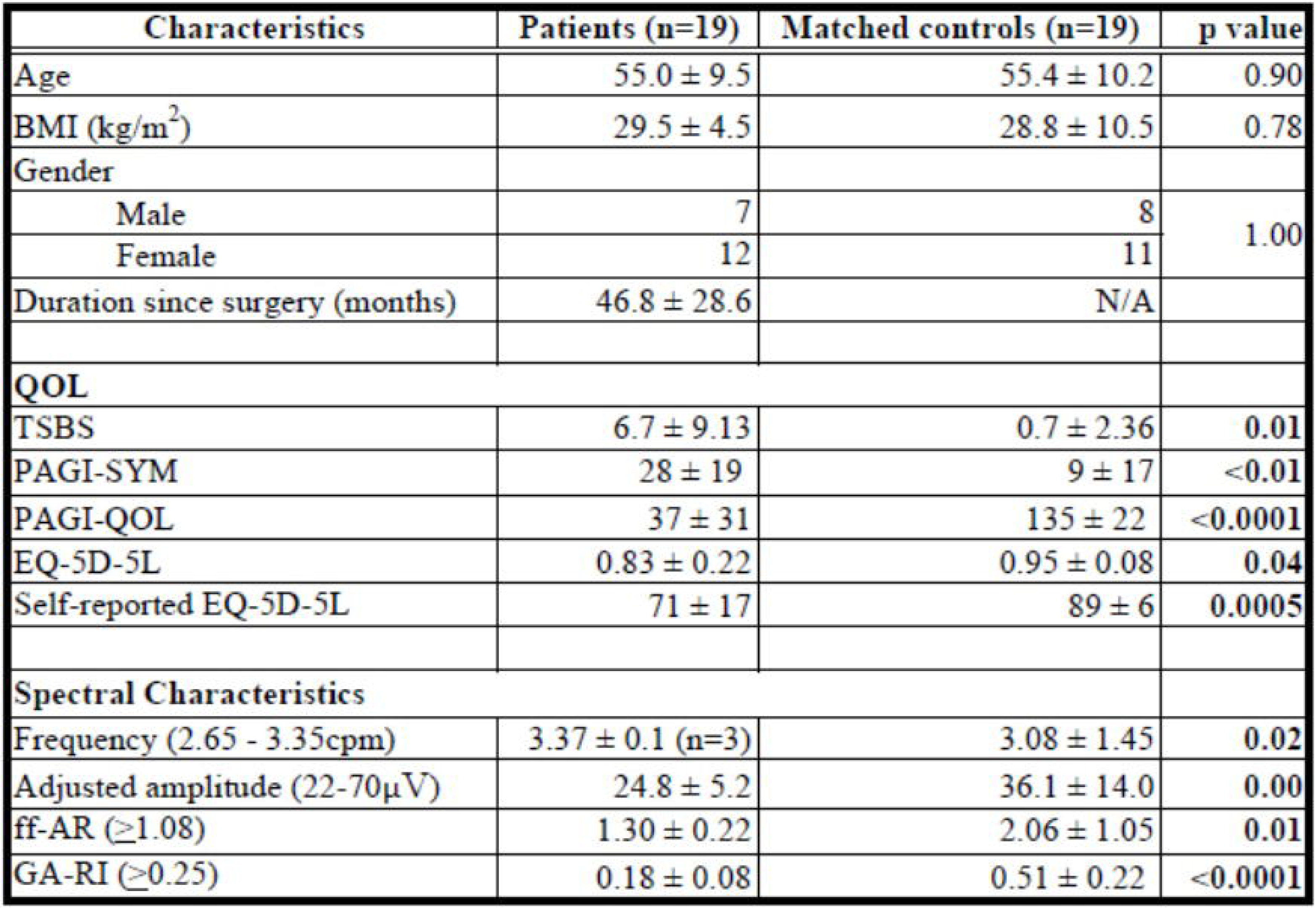

### Symptom Burden and Quality of Life

Differences in the Total Symptom Burden Score (TSBS), upper gastrointestinal symptoms and QoL scores between patients and controls are reported in **Table 1**, with patients showing substantially worse scores across all tests, including time-of-meal symptoms, PAGI-SYM and PAGI-QOL. Comparisons in QoL data and TSBS for the different bypass groups are presented in **Table 2**. The conversion to bypass group had a lower average QoL compared to the bypass-only group, however, this did not reach statistical significance within the present cohort size (6 vs 13).

| Spectral characteristics | Bypass (n=13) | Conversions (n=6) | p value | Overall (n=19) |
| --- | --- | --- | --- | --- |
| Frequency (2.65 - 3.35) | N/A | N/A | N/A | $3.37 \pm 0.1$ |
| Adjusted amplitude (22-70) | $25.0 \pm 3.8$ | $24.3 \pm 8.4$ | 0.86 | $24.8 \pm 5.2$ |
| ff-AR ($\geq 1.08$) | $1.28 \pm 0.22$ | $1.34 \pm 0.22$ | 0.57 | $1.30 \pm 0.22$ |
| GA-RI ($\geq 0.25$) | $0.20 \pm 0.08$ | $0.13 \pm 0.06$ | 0.07 | $0.18 \pm 0.08$ |
| TSBS (Symptom Score $\geq 3$) | 62% (8/13) | 83% (5/6) | | 68% (13/19) |

### Gastric Spectral Data

18/19 participating patients had at least 1 abnormal gastric slow wave spectral abnormality in the remnant stomach, including 17 patients who had 2 or more abnormalities compared to the test reference intervals (25). Data for all spectral metrics by group are presented in **Table 3**. Due to the very low GA-RI value for almost all patients (17/19), the Principal Gastric Frequency values could only be identified in 3 subjects (2 of the 3 having normal GA-RI values). By contrast, a Principal Gastric Frequency was reliably detected in all controls, including during their fasting periods. All 3 cases with a preserved PGF were in the bypass-only group, with an average time since bypass of 38.8 ± 25.1 months. The GA-RI value was lower in the conversion-only group vs. the bypass-only group, although this did not reach significance (0.020 ± 0.08 vs 0.13 ± 0.06; p=0.07) (**Table 3**).

| QOL | Bypass (n=13) | Conversions (n=6) | p value | Overall (n=19) |
| --- | --- | --- | --- | --- |
| TSBS | 3.8 ± 3.4 | 12.9 ± 14.3 | 0.18 | 6.7 ± 9.13 |
| PAGI-SYM | 22 ± 11 | 41 ± 26 | 0.14 | 28 ± 19 |
| PAGI-QOL | 30 ± 24 | 52 ± 40 | 0.26 | 37 ± 31 |
| EQ-5D-5L | 0.85 ± 0.24 | 0.80 ± 0.19 | 0.67 | 0.83 ± 0.22 |
| Self-reported EQ-5D-5L | 74 ± 16 | 65 ± 19 | 0.30 | 71 ± 17 |

A reference spectral map of a typical healthy control is presented in **Figure 1D**, together with the Gastric Alimetry test reference intervals. Example spectrograms from a bypass case and conversion to bypass case, with highly degraded gastric activity by comparison, are shown in **Figure 2A and B** respectively. Summary spectrograms (combining all subjects’ data) are shown for the bypass-only group, conversion to bypass group, and controls, in **Figures 2C-E**, again showing dramatically degraded gastric activity following bypass. Comparisons to reference intervals are depicted in **Figure 2F**, with the main abnormality being reduced GA-RI.

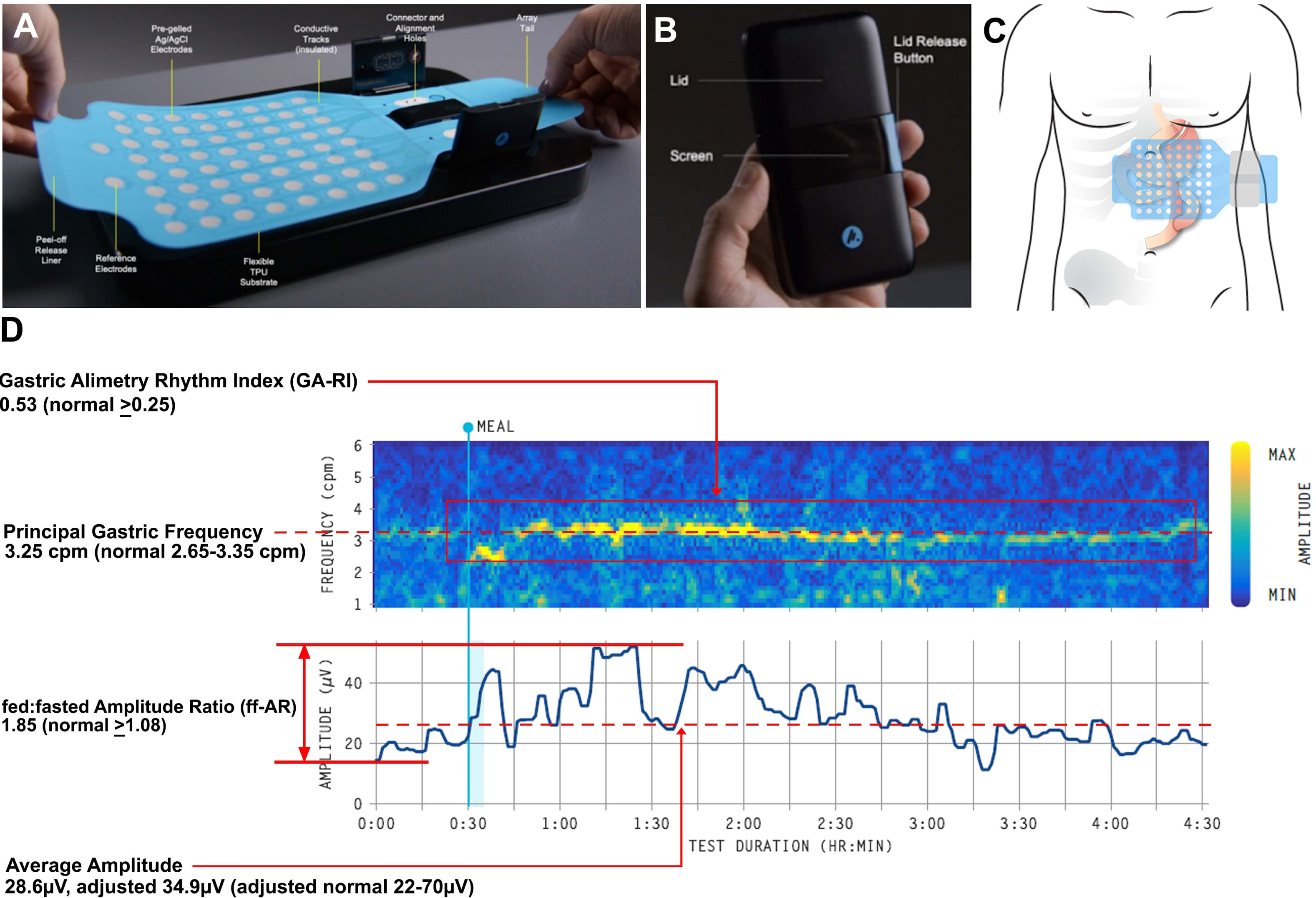

### Correlational Analysis

Higher PGF (r=-0.57), lower ff-AR (r=0.42), lower GA-RI (r=0.63), and lower BMI-adjusted amplitude (r=-0.43) were all strongly associated with lower quality of life as measured by the PAGI-QOL score (p<0.05). Similarly, lower GA-RI (r=-0.41), and lower BMI-adjusted amplitude (r=-0.38) were also significantly associated with higher symptom burden as measured by PAGI-SYM score (p<0.05). Selected correlation plots for BMI-adjusted amplitude and GA-RI are shown in **Table 3**. For specific symptoms, PGF was found to be correlated with early satiation, BMI-adjusted amplitude was correlated with nausea and vomiting, and ff-AR was correlated with early satiation and symptoms of heartburn and regurgitation (p<0.05).

### Safety

No adverse events were identified in any subject.

## Discussion

Gastric bypass and conversion to gastric bypasses involve reconstruction of the gastrointestinal tract with exclusion of a gastric remnant. Post-operative symptoms can be persistent, including nausea, vomiting, pain and dumping, however, the pathophysiology may be unclear despite normal endoscopy or imaging (2). The results of this study show that over one third of patients experienced moderate to severe symptoms with worse QoL compared to matched controls. Remnant gastric stomach function was also found to be highly degraded in the patient cohort, with many of these changes found to be correlated to worse symptom burdens and lower quality of life scores.

The associations between remnant stomach degeneration, foregut symptoms and quality of life in this study are novel. Regarding the degraded gastric pacemaker system in the bypassed stomach, this is expected due to disuse degeneration, as pacemaker system degeneration has also been described in animal stomachs after marked calorie restriction, thought to be mediated in-part by reduced local production of IGF-1 (26). Gastric bypass in itself could also conceivably modify the gastric electrical conduction system through disconnection of the stomach and partial denervation (27). Regarding the symptom associations, it is plausible that the gastric dysrhythmias could contribute to upper gastrointestinal symptoms in bypass patients, as post-surgical gastric dysrhythmias have been linked to symptoms in some other post-surgical populations previously, although causation was not assessed in the current study (14, 15, 28, 29).

Bypass patients can experience significant long-term symptoms and QOL impairment compared to matched controls, particularly in the group undergoing conversion to bypass. While these findings were not unexpected, given these patients have undergone surgery and typically have comorbidities, the findings further quantify the long-term symptom burdens and QoL of patients after bypass procedures, with one-third suffering moderate to severe post-prandial symptoms in this series. Such outcomes should be understood by surgeons and patients embarking on these procedures, while noting that disease indications for surgery may also significant impact prior quality of life.

The GA-RI was reduced in the conversion group compared to the bypass-only group, which neared significance (p=0.07) even despite the small number of conversion patients in this subgroup (n=6). This is likely a true finding, reflecting the effect of the previous vertical sleeve gastrectomy, which removes the native pacemaker and will have already caused underlying gastric rhythm disturbances (14). The adjusted amplitude and the ff-AR reductions compared to the matched control cohort, are explained by pacemaker degeneration, smooth muscle atrophy, a reduced volume of electrically active functioning gastric tissue and the lack of food content stimulating the gastric remnant.

While there have been previous studies assessing the effects of surgical techniques on the post-operative outcomes and quality of life in bypass, along with manometric assessments of the gastric pouch and the Roux limb, studies of the remnant gastric conduction system have been sparse prior to this study (30, 31). This is likely due to the lack of adequately high-resolution techniques and devices to assess the electrophysiology of the stomach. Recent technological advances in the form of invasive and non-invasive high-resolution gastric mapping techniques have now significantly improved our understanding of gastric electrophysiology in health and disease (9, 32, 33). Legacy techniques including electrogastrography (EGG) have previously attempted to assess the electrical activity of the stomach following surgery, however, this is limited by low resolution and high sensitivity to noise. Gastric Alimetry overcomes these problems by employing an HR array together with sophisticated signal processing algorithms (22, 23), to non-invasively assess the electrophysiology of the gastric remnant and to also simultaneously record symptom development and progression. Previously, Gastric Alimetry has been exclusively performed in patients with normal gastric anatomies, in whom reference ranges were developed (24, 25, 33). As this study now involves a modified Gastric Alimetry protocol to account for the effect of surgery on the stomach, the authors used both the validated reference range and a cohort of matched control patients (with a similar calorie load meal challenge) as a comparison.

The strengths of this study include the recruitment of symptomatic and asymptomatic patients in the patient population, and the use of a matched control cohort for comparison. Robust statistical analyses were also performed to assess for differences between these cohorts. The main limitation of this study was its dominant focus on remnant gastric motility, such that other potential sources of symptoms such as pouch and Roux-limb function, together with other contributing disorders, were not systematically evaluated. A causal association should not be inferred from our results, particularly because the remnant stomach is excluded from GI continuity and symptoms had a post-prandial character. Therefore, it is highly likely that symptom genesis post-bypass is multi-factorial, with other contributing sources including gastric pouch stretch, Roux stasis syndrome, and post-surgical sequelae (31, 34). Future work could now focus on addressing the causality of the novel finding that degradations of gastric electrophysiology correlated with symptoms, which could be approached using frameworks such as the Plausibility Criteria proposed by Tack et al (35).

The results of this study may have clinical implications beyond sleeve gastrectomy. In particular, the finding that a diverted stomach develops severe degeneration of the gastric conduction apparatus may become relevant in the pathophysiology and management of other medical or surgical conditions where there is prolonged stomach disuse. Relevant conditions include prolonged illness with caloric restriction, including patients on long-term nasojejunal feeding, total parenteral nutrition, and severe anorexia. When clinically rehabilitating these patients back onto normal oral diets after a period of gut atrophy, clinicians should be cognizant that stomach pacemaker function may have become degraded, requiring additional time for recovery with return of dietary tolerance. Recovery patterns may also be variable, due to the inherent recovery capacity of the pacemaking, enteric neural and smooth muscle components (36, 37), however, this requires further study.

In conclusion, a third of gastric bypass patients experienced significant long-term upper GI symptoms and a reduced quality of life. These consequences are associated with severe degradation of remnant gastric electrophysiological function, although overall symptom genesis in post-bypass patients is more likely to be multifactorial.

## Data Availability

All data produced in the present study are available upon reasonable request to the authors

## Acknowledgements

We thank the volunteers who participated in this research, the Auckland City Hospital clinical nurse specialist Elaine Yi and our Auckland clinical research coordinators Gen Johnston and India Wallace.

